# Pattern, treatment modalities and radiological outcome of pediatric femoral shaft fractures treated in Northern,Tanzania

**DOI:** 10.1101/2024.03.07.24303832

**Authors:** Shindo I. Kilawa, Anthony J. Pallangyo, Elifuraha G. Maya, Rogers J. Temu, Faiton N. Mandari, Frank I. Olotu, Estomick K. Ofunguo, Adnan M. Sadiq, Honest H. Massawe, Octavian Shirima, Reginald Shoo

**Author notes:** Corresponding author: Shindo Isack Kilawa, Email addresses (SIK). These authors contributed equally to this work (Shindo I. Kilawa, Anthony J. Pallangyo, Rogers J. Temu, Frank I. Olotu, Reginald Shoo). (EGM). (AJP). (RJT). (RS). (FNM). (HHM). (FIO). (AMS). (EKO). (OAS).

## Abstract

**Background:** Femoral shaft fracture is among the most common causes of paediatric hospitalisation, mortality and morbidity worldwide. There is no clear option that is preferable to other treatment modalities, especially between 5 to 16 years and published studies are scarce on radiological outcomes in Sub-Saharan Africa.This study aimed to determine the pattern, treatment modalities and radiological outcome of the paediatric femoral fractures treated at KCMC.

**Methodology:** A cross-sectional study was conducted for all children with femoral shaft fractures treated at Kilimanjaro Christian Medical Centre from 1^st^ January 2018 to 31^st^ December 2022. The approval to conduct the research was obtained from Kilimanjaro College Research Ethics and Review Committee (CRERC) with ethical clearance Reg NO PG 88/2022. In our study we used secondary data and the permission to conduct the research was obtained from KCMC, hence no formal consent was required from patients/parents.The data was accessed from the files and Elecronic Health Management system (EHMS) from 01/04/2023 to 31/07/2023.The radiological outcome; shortening, angulations in six weeks and fracture union, 12 weeks post-management were reviewed with the involvement of a consultant radiologist and the orthopaedic surgeon to obtain the precise information and were recorded on the extraction sheet.

**Results:** This study included 230 study participants who met the inclusion criteria. The mean age of participants was 9.1 (5.1) years, 41.3%, was aged 6 – 12 years, 82.2% were males, 45.7% were involved in a MTC, and 83.5% had no health insurance. The commonest fracture type was 92.6% closed, 48.7% transverse, and 65.% right side. Non-operatively was used in 50.9% of which 76.8% were treated with late hip Spica. Of those treated operatively, 61.1% were plating.

The majority had good radiological outcomes with acceptable solid union, angulation and shortening. Those patients who were not operated had 94% lower odds of satisfactory radiological outcomes than those who were operated ( AOR=0.06, 95% CI: 0.01 – 0.27 and p<0.001) whereas other factors were not statistically significant.

**Conclusion:** The majority of the study participants were male and were involved in MTC as the commonest mechanism of injury. Most had closed fractures that mainly presented on the right side and transverse fractures were the most common type. The hip Spica was common non-operatively option; however, plating was the most common operative option. Treatment modality substantially affected radiological outcomes and was statistically significant.

## Introduction

PFSF is among the most common causes of paediatric hospitalisation, morbidity, and mortality worldwide (1,2). It is relatively uncommon as compared to the adult population, although it’s occurrence poses a high burden in morbidity in terms of cost and time of treatment(3). Paediatric femoral shaft fracture affects children’s life individually, socially, educationally and emotionally, (4) and also can contribute to long-term disability (5).

Femoral shaft fractures in paediatrics account for about 1.6% of all paediatric fractures globally (6–8). Paediatric femoral shaft fractures commonly affect children aged 2 to 12 years (5) with boys more commonly affected than girls (7,9).

Falls and motor traffic crushes are the most common mechanism of injury (10,11) but depend on the child’s age (9). Falls were the most common mechanism of injury in younger children, while motor traffic accidents were the most common in older children and adolescents (8).

Treatment of PFSF aims to restore limb function and return to normal activities as soon as possible(12). There is an increasing trend towards operative modalities with increased technologies (3). There is no clear option that is preferable to other treatment modalities, especially between 5 to 16 years, although several studies recommend the use of flexible or Elastic nails (13,14).

Regardless of several modalities of treatment, complications including malunion is common, especially for non-operative treatment like hip spica (15).

Generally, a scarcity of published studies on radiological outcomes has been observed, especially in Sub-Saharan Africa, henceforth this study was conducted to determine the patterns, treatment modalities, and radiological outcomes of paediatric femoral fractures treated at KCMC.

## Material and methodology

### Study design and setting

This hospital-based cross-sectional study was conducted at Kilimanjaro Christian Medical Centre in Moshi district-kilimanjaro, North-eastern Tanzania from January 2018 to December 2022.

KCMC is among the largest referral teaching Hospitals in Tanzania owned by the Institution of Good Samaritan Foundation with a capacity of 696 normal beds plus 145 canvas making a total of 841 beds, serving patients from all regions of the northern part and some from the central part of Tanzania as well nearby countries.

### Study population

The study population was all paediatric patients with femur fractures who were treated at Kilimanjaro Christian Medical Centre during the study period.

### Inclusion criteria and exclusion criteria

This study recruited all paediatric patients with femoral shaft fractures treated at KCMC from 1^st^ January 2018 to 31^st^ December 2022 and excluded those with pathological fractures, infected open wounds, Neuromuscular disorders, missing information, and who died before discharge.

### Sample size and sampling technique

The minimum sample size was 225 patients calculated using the single proportion with an assumption of 18% prevalence obtained from the study done at KCMC (16); however, this study enrolled 230 study participants.

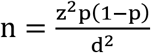

Where P= 0.18, Z =1.96 and d =0.05

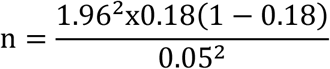

Therefore the estimated sample size (n) was 225; this study enrolled 230 study participants.

A non-probability, convenient sampling technique was used, hence all paediatric patients with femoral shaft fractures treated at KCMC and registered in admission and trauma registry books during the study period were included in the study.

### Study variables

Radiological outcome was the dependent variable whereas age, sex, residence, location at the time of injury, mechanism of injury, patterns of fracture and treatment modalities were used as independent variables.

### Data collection tools, methods and procedures

The data collection tools used were the structured extraction sheet and a Modified RUST score(reference Appendix 1). The data was accessed from the files and Elecronic Health Management system (EHMS) from 01/04/2023 to 31/07/2023. We identified all children admitted and registered in the admission books in the orthopaedic department. Their identification numbers were used to trace for their initial x-rays, control x-rays and follow-up x-rays of at least ≥ 6 weeks.

Those patients with available x-rays in radiological systems (OHIF/DICOM) and hard copies with consideration of inclusion and exclusion criteria were traced and reviewed to obtain demographic information, clinical characteristics and mode of treatment from the EHMS and medical records files.

The principal investigator and assistant researchers reviewed the initial and follow-up x-rays with the involvement of a consultant radiologist and the orthopaedic surgeon to obtain precise information regarding the pattern of fracture, classification of fractures and radiological outcome that were recorded on a structured extraction sheet.

The anterior-posterior and lateral view x-ray of the full-length femur after bridging callus formation before full healing and remodelling were used to measure shortening and angulation by using the measurement in the system, ruler and goniometer (protractor) for the hard copies x-rays.

Based on previous studies, we defined malunion as a healed fracture with a shortening of greater than 2cm and greater than 15 degrees recurvatum or procurvatum or greater than 10 degrees valgus or varus (17).

We assessed the union rate after three months using a modified RUST score; a score of 11 to 16 was considered a reasonable union rate.

Satisfactory radiological outcomes refer to all acceptable parameters. Therefore, if any of the parameters above are unacceptable, the outcome were considered unsatisfactory.

### Data management and analysis plan

The obtained information was documented on the structured extraction sheets daily basis and cross-checked to ensure completeness and minimise errors before data analysis.

The complete data were entered, processed and analysed using SPSS version 25. Cleaned data, and new variables were created and categorised where necessary. Descriptive statistics were summarised using frequency and proportion for categorical variables and measures of central tendency with respective measures of dispersion for continuous variables.

Fisher’s exact test establishes a relationship and comparison between treatment options, radiological outcomes and explanatory variables.

Logistic regression was performed to obtain an odds ratio (ORs) with a 95% confidence interval (CI) for the association between a set of explanatory variables and radiological outcomes. Those with a p-value less than 0.05 were considered statistically significant.

### Ethical consideration

The approval to conduct the research was obtained from Kilimanjaro College Research Ethics and Review Committee(CRERC) with ethical clearance Reg NO PG 88/2022. In our study we used secondary data and the permission to conduct the research was obtained from Kilimanjaro Christian Medical Center adminstration hence no formal informed consent was required as we didn’t meet with patients during the data collection. No patient identifying information such as file numbers and patient names were used insteady we used coding numbers.

## RESULTS

### Demographic characteristics of the study participants

This study included a total of 230 study participants. The study participant’s mean (SD) age was 9.1 (5.1) years. The majority of the study participants, 95 (41.3%), were aged 6 – 12 years, 189 (82.2%) were males, 155 (67.4%) were residing in rural areas,119(51.7%) were on the street at the time of injury, 192 (83.5%) had no health insurance support. Some children had associated injuries.—This is summarised in Table 2.

**Table 1:** Satisfaction radiological outcomes parameter.

| Parameters | Acceptable or satisfactory |
| --- | --- |
| Anterior or posterior angulation | $\leq 15$ degrees |
| Valgus or Varus | $\leq 10$ degrees |
| Shortening | $\leq 2$ cm |
| Modified RUST Score | 11 to 16 |

**Table 2:**
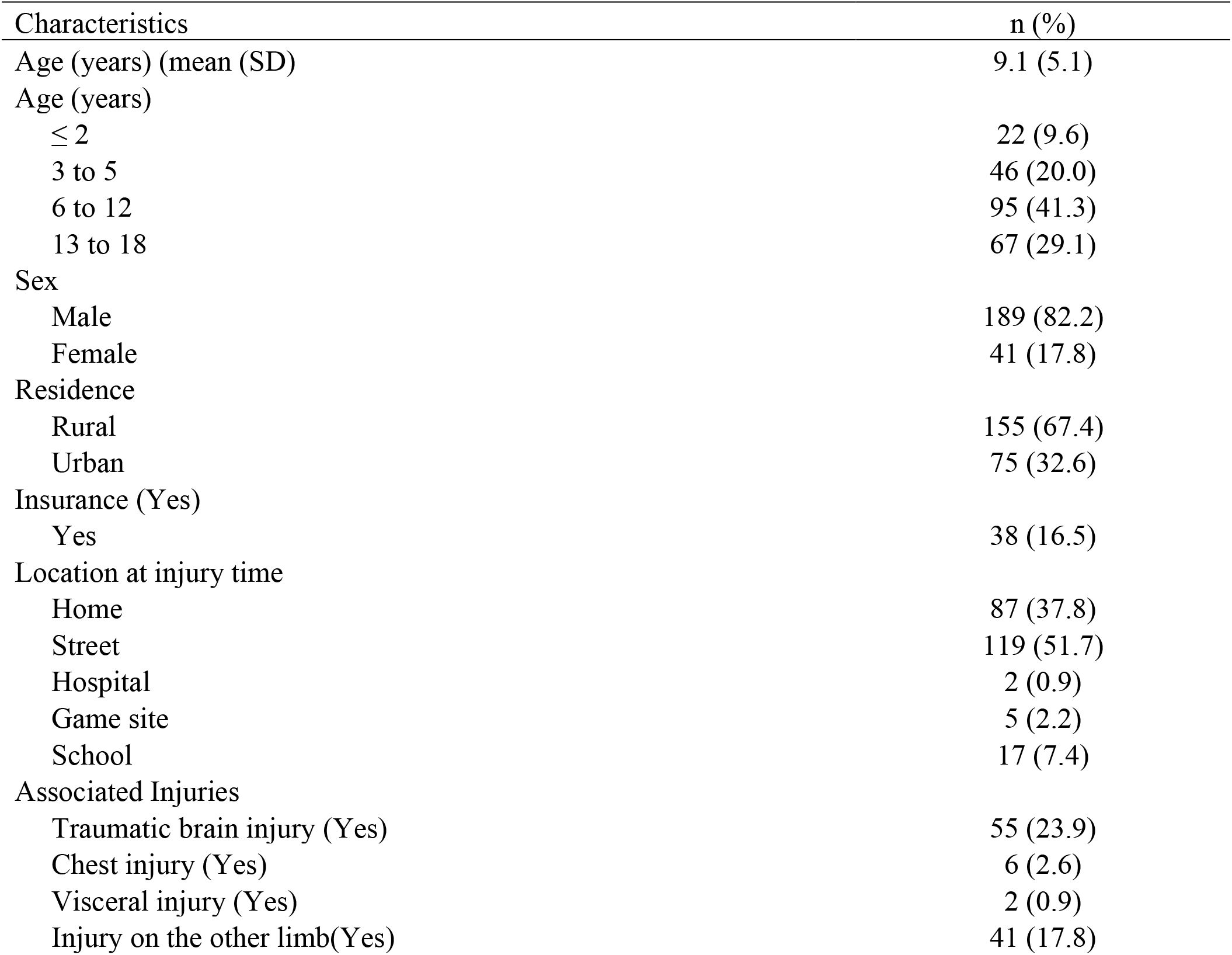
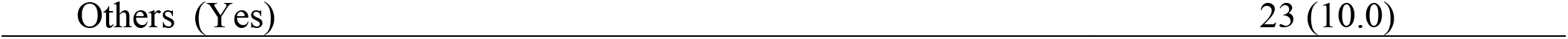
Demographic and clinical characteristics of the study participants (n=230)

### The pattern of the paediatric femoral shaft fractures

The pattern of paediatric femoral fracture was as follows; 128 (55.6%) had injured the right-side limbs, 213 (92.6%) sustained closed injuries, 150 (65.2%) had mid-shaft fractures, 112 (48.7%) had transverse fractures, 105 (45.7%) were involved in motor vehicle crash, 119 (51.7%) Refer to Table 3.

**Table 3:** The pattern of the paediatric femoral shaft fractures treated at KCMC (n=230)

| Fracture pattern | n (%) |
| --- | --- |
| Injured side |  |
| Right | 128 (55.7) |
| Left | 102 (44.3) |
| Soft tissue status |  |
| Closed | 213 (92.6) |
| Open | 17 (7.4) |
| Fracture location |  |
| Proximal | 37 (16.1) |
| Middle | 150 (65.2) |
| Distal | 43 (18.7) |
| Types of fracture |  |
| Transverse | 112 (48.7) |
| Comminuted | 33 (14.4) |
| Spiral | 28 (12.2) |
| Oblique | 54 (23.5) |
| Segment | 3 (1.3) |
| Mechanism of injury |  |
| Motor traffic crush | 105 (45.7) |
| Fall | 86 (37.4) |
| Non-accident trauma/Assault | 7 (3.0) |
| Heavy objects fall on | 24 (10.4) |
| Birth injury | 2 (0.9) |
| Bike accident | 6 (2.6) |

### The treatment modalities of paediatric femoral shaft fractures

Approximately half of the study participants, 117 (50.9%), were treated non-operatively, and the remaining treated operatively. The majority of those treated non-operatively, 90 (76.8%), were treated by late hip spica. The majority of those treated operatively, 69 (61.1%), were treated by plating.—This is summarised in figures 1 & 2.

**Figure 1:** Non-operative treatment options (n= 117)

**Figure 2:** Operative treatment options (n=113)

**Figure 3:** Showing x-rays of a child, treated with flexible nails,(a) Preoperative, (b)immediately postoperative, (c)&(d) 6 weeks follow-up-From Digital Imaging and Communication in Medicine.

**Figure 4:** Showing x-rays of child, treated with late hip spica,(a) X-rays on admission and (b) 6-week follow-up X-ray, From Digital Imaging and Communication in Medicine.

### The radiological outcome of the paediatric femoral shaft fractures

Most of our study participants had good radiological outcomes with the solid union in 100%, acceptable angulation and shortening. However, the minority had malunion in which 0.4% had valgus, 13% had varus of more than 10 degrees, 6.1% had anterior angulation of more than 15 degrees and none had unacceptable posterior angulation. Only 3.1 % had limb shortening of more than 2cm, as shown summarised in Table 4

**Table 4:** The radiological outcome of the paediatric femoral shaft fractures treated at KCMC.

| Radiological outcome | n (%) |
| --- | --- |
| Limb shortening (cm) |  |
| $\leq 2$ | 223 (96.9) |
| $> 2$ | 7 (3.1) |
| Coronal angulation in degrees |  |
| Valgus |  |
| $\leq 10$ | 229 (99.6) |
| $> 10$ | 1(0.4) |
| Varus |  |
| $\leq 10$ | 199(86.5) |
| $> 10$ | 31(13.5) |
| Sagittal angulation in degrees |  |
| Anterior |  |
| $\leq 15$ | 216(93.9) |
| $> 15$ | 14 (6.1) |
| Posterior |  |
| $\leq 15$ | 230(100.0) |
| $> 15$ | 0(0.0) |

**Table 5:**
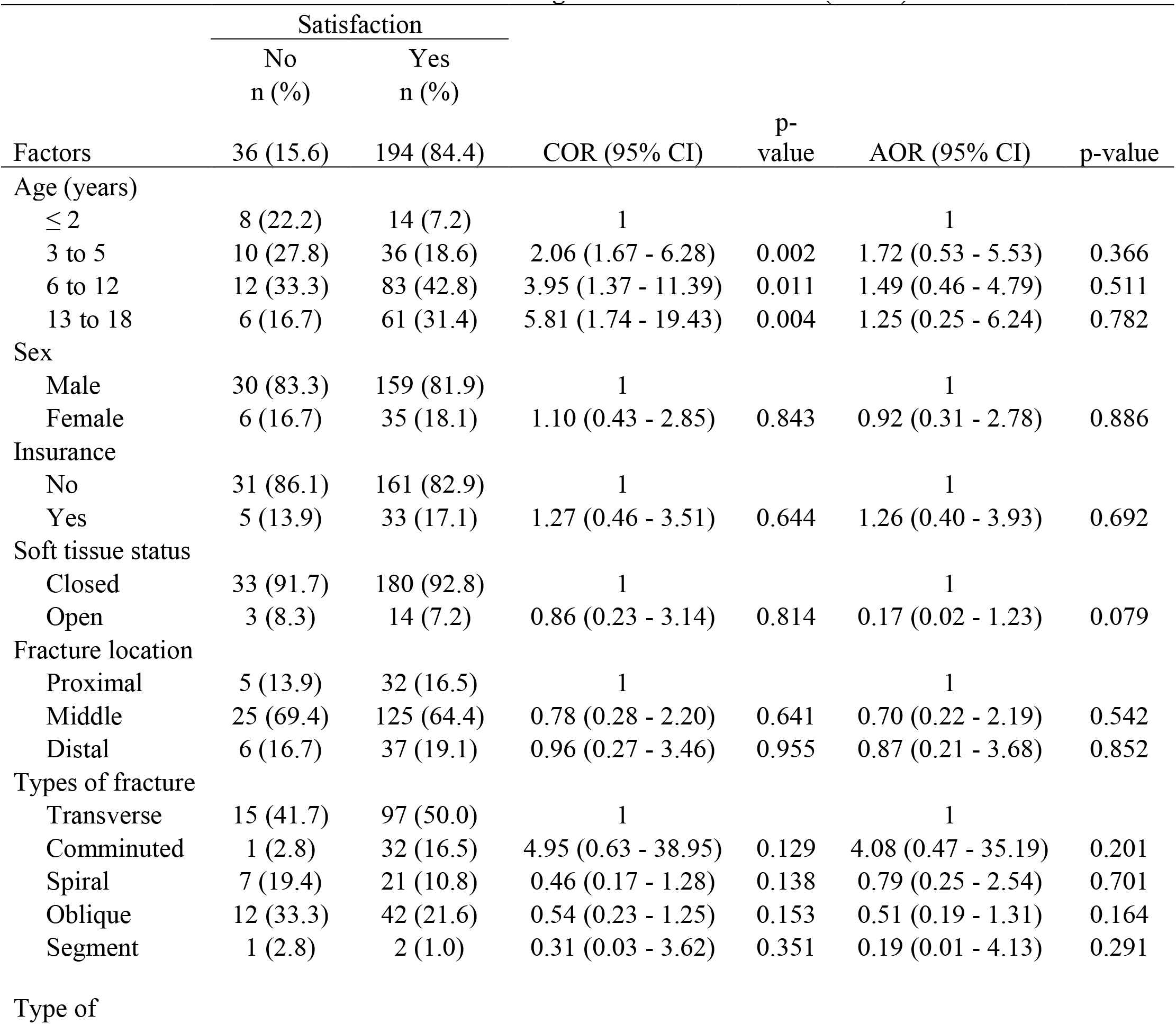
Factors associated with radiological outcomes of PFSF (n=230)

### Factors associated with the radiological outcome of the PFSF

Those managed non-operatively had 94% lower odds of satisfactory radiological outcomes than those operated after adjustment of confounders, which was statistically significant, as shown in Table.

## Discussion

This study found that the mean age of the study participants was 9.1(5.1 years), with male predominance and more than three-quarters had no health insurance for support during management. Regarding treatment, in this study, non-operative treatment was slightly preferable to approximately more than half of the study participants of which late hip Spica was the most common of all non-operative modalities.

The current practice encourages surgical intervention more than non-operative treatment, especially in the United Kingdom. These modalities allow early mobilization and shorter hospital stays (3,12). However, all modalities had a good outcome (11).

In our study, almost all participants had good radiological outcomes with solid union in 100%. Nevertheless, those who were managed non-operatively had 94% lower odds of satisfactory radiological outcomes than those who were operated on.

### The pattern of the paediatric femoral shaft fractures

#### Mechanism of Injury

This study revealed that the leading mechanism of injury was MTC, relatively similar to the studies done in India and Nigeria (3,15), hence the community has the responsibility to protect children from MTC while are on the street, more than half of the study participants sustained an injury on the street, that can be explained due to significant increase in motorcycles (Motorbike and motor vehicles) as means of transportation in our country. In contrast with the studies done in Singapore and Cameroon where falls were reported as a leading mechanism of injury (9,18).

However, the mechanism of injury depends on the child’s age (8,9). Falls were the leading mechanism of injury in younger children, while motor traffic accidents were the most common in older children and adolescents (8).

#### The Location of Fracture and Type of Fracture

In this study, the mid-shaft was the most common location of the fracture similarly observed in the study done by Hoffman et al and Guifo et al (9,19) and the majority had a closed fracture, similarly reported in Nigeria, and Tanzania (3,16).

Transverse followed by an oblique fracture was the most common of fracture as similarly reported by other studies done in Singapore and Nepal (14,18). This is due to the direct impact on the femur due to MTC as the most common mechanism of injury. This was observed differently in the study by Rush et al, where spirals followed by transverse were the most common, the study involved only 10 participants younger than one year old (20).

#### The treatment modalities of paediatric femoral shaft fractures

The mode of treatment of paediatric femur fracture remains in debate in several studies, there is no ideal modality as in adults. In this study, non-operative treatment was commonly used in managing femoral shaft fractures in children, accounting for 50.9%, and mostly were on late hip Spica. Similar observations were seen in other studies (14,16,21)

Several studies favour the use of flexible nails at ages 6 to 12 years. Flexible nails can prevent fracture rotation and maintain the length and alignment at the fracture site depending on fracture pattern (14). However, in our study, only 2.7% were managed with flexible nails and all had good radiological outcomes. There is a need to adopt the use of flexible nails in our setting regardless of minimal differences in outcome

#### The radiological outcome of the paediatric femoral shaft fractures

In our study, almost all study participants had good radiological outcomes with a solid union in 100% and the majority had acceptable angulation and shortening, similarly as reported in other studies (3,9,10,14,18,20). The good radiological outcome after femoral shaft management is generally explained by the potential ability to remodel in paediatric regardless of malaligned fracture.

This study found that treatment options were statistically significantly associated with satisfaction with radiological outcomes while other factors were not. Those who were managed non-operatively had 94% lower odds of satisfactory radiological outcomes than those operated, which favour operative management similarly as in other studies, however, they found age has a strong effect on the outcome (10). Differently, a randomized controlled trial done by Shemshakij in Iran reported treatment options were not statistically significantly associated with radiological outcomes (22).

#### Study Limitations and Strengths

##### The Study limitations

This was a single-centre study, done retrospectively,with too small numbers of different ages and too short follow-up time, hence encountered incompleteness of patient information, incredibly initial, post-definitive treatment and follow-up x-rays.

##### The study strength

The sample size was adequate to meet the goal as expected after sample size calculation, baseline information was obtained well and observed confidentiality.

## Conclusion

The majority of the study participants were male and involved in MTC as the leading mechanism of injury. Most of the study participants had closed fractures that mainly presented on the right side, in which transverse fractures were the leading type, followed by oblique fractures.

Non-operative was preferable to operative treatment, and the hip spica was the most common in non-operative modalities; however, the plating was the most common operative option. There was not much difference in radiological outcome regardless of the modality of treatment used; however, Treatment options were statistically significantly associated with satisfaction with radiological outcomes.

## Recommendations

Proper implementation of road traffic policies to reduce MTC injuries should be addressed, as the study found that motor traffic crashes are a significant cause of injury.

In the future, prospective or multicenter-based study studies should be done to compare treatment modalities used and radiological outcomes that will involve all children as others might have been treated at other facilities and bone settlers.

## Data Availability

The data is available at the institution, not shared online due to the ethical reasons

## Acknowledgements

I am grateful to the almighty God who protected me throughout my life, especially during my Orthopedic and Traumatology studies training. I sincerely thank Tumaini University, and Kilimanjaro Christian Medical University College for their acceptance and assistance in my daily studies.

I also thank the Department of Radiology, Registry and ICT for supporting my research activities. I want to thank my fellow residents, nurses, medical attendants, intern doctors, and medical doctor students for their tireless support and input.

I also thank my lovely sons Isaac and Collins and my lovely wife Miss Irene, who had to suffer his father’s absence during my studies. I also thank my lovely father, Mr Isack Kilawa, and my beloved late mother Mrs Luciana Isack, who raised and gave support since my birth. I want to thank my Research Assistants, Dr Irene Tendwa and Irene Rabieth. I will be selfish of good if I don’t extend my gratitude to all who ever supported me during my training, either directly or indirectly.

## Appendix I A modified RUST score tool for assessment of fracture union

| Score per Cortex | Callus | Fracture Line |
| --- | --- | --- |
| 1 | Absent | Visible |
| 2 | Present | Visible |
| 3 | Bridging | Visible |
| 4 | Remodelled | Invisible |

A score done to each (anterior, posterior, medial, lateral) RUST score is the sum of all cortex scores, with four as a minimum and 16 as a maximum score (Litrenta et al.,2015).

## Appendix II Ethical Clearance

## References

1. WHO. INJURIES VIOLENCE THE FACTS The magnitude and causes of injuries. Geneva World Heal Organ [Internet]. 2014;20. Available from: https://www.who.int/healthinfo/global_burden_disease/projections/en/

2. Lodamo T, Worku A, Desta T, Shimelis T, Wondwossen Elssa N. PATTERN OF PEDIATRIC FEMUR SHAFT FRACTURES IN A TERTIARY HOSPITAL, ADDIS ABABA, ETHIOPIA. Vol. 58, Ethiop Med J. 2020.

3. SE I, J C. Femoral Fractures in Children Treated in a Regional Trauma Center in Nigeria. Int J Crit Care Emerg Med [Internet]. 2019 Mar 14;5(2). Available from: https://www.clinmedjournals.org/articles/ijccem/international-journal-of-critical-care-and-emergency-medicine-ijccem-5-070.php?jid=ijccem

4. Kumar R, Kumari A, Pandit A. Evaluation of outcome of titanium elastic nailing (TEN) versus hip spica cast in the treatment of femoral shaft fractures in children. Int J Orthop Sci [Internet]. 2018 Oct 1;4(4):149–54. Available from: http://www.orthopaper.com/archives/?year=2018&vol=4&issue=4&ArticleId=1138

5. Madhuri V, Dutt V, Gahukamble AD, Tharyan P. Interventions for treating femoral shaft fractures in children and adolescents. Evidence-Based Child Heal A Cochrane Rev J [Internet]. 2014 Dec 15;9(4):753–826. Available from: 10.1002/ebch.1987

6. Tisherman RT, Hoellwarth JS, Mendelson SA. Systematic review of spica casting for the treatment of paediatric diaphyseal femur fractures. J Child Orthop [Internet]. 2018 Apr 1;12(2):136–44. Available from: 10.1302/1863-2548.12.170201

7. Akinyoola A, Orekha O, Taiwo F, Odunsi A. Outcome of non-operative management of femoral shaft fractures in children. African J Paediatr Surg [Internet]. 2011 Jan;8(1):34. Available from: http://www.afrjpaedsurg.org/text.asp?2011/8/1/34/78666

8. Engström Z, Wolf O, Hailer YD. Epidemiology of pediatric femur fractures in children: the Swedish Fracture Register. BMC Musculoskelet Disord [Internet]. 2020 Dec 1;21(1):796. Available from: 10.1186/s12891-020-03796-z

9. Guifo ML, Tochie JN, Oumarou BN, Moulion JR, Bang AG, Ndoumbe A, et al. Paediatric fractures in a sub-saharan tertiary care center: a cohort analysis of demographic characteristics, clinical presentation, therapeutic patterns and outcomes. Pan Afr Med J [Internet]. 2017 May 18;27. Available from: http://www.panafrican-med-journal.com/content/article/27/46/full/

10. Sela Y, Hershkovich O, Sher-Lurie N, Schindler A, Givon U. Pediatric femoral shaft fractures: treatment strategies according to age - 13 years of experience in one medical center. J Orthop Surg Res [Internet]. 2013 Dec 17;8(1):23. Available from: 10.1186/1749-799X-8-23

11. Vitiello R, Lillo M, Donati F, Masci G, Noia G, Santis V De, et al. Locking plate fixation in pediatric femur fracture: Evaluation of the outcomes in our experience. Acta Biomed [Internet]. 2019;90(4):110–5. Available from: http://www.ncbi.nlm.nih.gov/pubmed/32934867

12. Khoriati A achraf, Jones C, Gelfer Y, Trompeter A. The management of paediatric diaphyseal femoral fractures: a modern approach. Strateg Trauma Limb Reconstr [Internet]. 2016 Aug 31;11(2):87–97. Available from: 10.1007/s11751-016-0258-2

13. Gyaneshwar T, Nitesh R, Sagar T, Pranav K, Rustagi N. Treatment of pediatric femoral shaft fractures by stainless steel and titanium elastic nail system: A randomized comparative trial. Chinese J Traumatol [Internet]. 2016 Aug 1;19(4):213–6. Available from: https://linkinghub.elsevier.com/retrieve/pii/S100812751630058X

14. Panthi S, Shrestha R, Pradhan J, Neupane B, Bhusal I, Karki A. Diaphyseal Femur Fracture in Paediatric Age group: Outcome with fixation by Elastic Nailing System. Eur J Med Sci [Internet]. 2021 Jan 27;3(1):1–5. Available from: https://www.europasianjournals.org/ejms/index.php/ejms/article/view/258

15. Singh Sandhu K, Kaur M, Singh H, Sandhu A. Evaluation of outcome, safety, and efficacy of diaphyseal fracture of femur and tibia in children-treated by Titanium Elastic Nailing System (TENS). Vol. 08, European Journal of Molecular & Clinical Medicine. Patiala-India; 2021.

16. Albert P, Honest M. Epidemiology and Associated Injuries in Pediatric Femoral Shaft Fracture Treated at a Limited Resource Zonal Referral Hospital in Northern Tanzania. Ann Orthop Musculoskelet Disord. 2021 Mar 15;4(1):1028.

17. E DT, C ER, E AF. Pattern and Treatment of Femoral Shaft Fracture in a Tertiary Hospital: One Year Retrospective Review. SAS J Surg. 2022 May 13;8(5):374–84.

18. Lee Y, Lim K, Gao G, Mahadev A, Lam K, Tan S, et al. Traction and Spica Casting for Closed Femoral Shaft Fractures in Children. J Orthop Surg [Internet]. 2007 Apr 4 [cited 2022 Apr 23];15(1):37–40. Available from: 10.1177/230949900701500109

19. Hoffmann CR, Traldi EF, Posser A. EPIDEMIOLOGICAL STUDY OF CHILDREN DIAPHYSEAL FEMORAL FRACTURES. Rev Bras Ortop (English Ed [Internet]. 2012 Mar;47(2):186–90. Available from: https://linkinghub.elsevier.com/retrieve/pii/S2255497115300847

20. Rush JK, Kelly DM, Sawyer JR, Beaty JH, Warner WC. Treatment of Pediatric Femur Fractures With the Pavlik Harness. J Pediatr Orthop [Internet]. 2013 Sep;33(6):614–7. Available from: https://www.pedorthopaedics.com

21. Ricardo Hoffmann C, Franceschini Traldi E, Posser A. EPIDEMIOLOGICAL STUDY OF CHILDREN DIAPHYSEAL FEMORAL FRACTURES [Internet]. Brazil; 2011 May. Available from: https://www.scielo.br/rbort

22. Shemshaki HR, Mousavi H, Salehi G, Eshaghi MA. Titanium elastic nailing versus hip spica cast in treatment of femoral-shaft fractures in children. J Orthop Traumatol [Internet]. 2011 Mar 22;12(1):45–8. Available from: 10.1007/s10195-011-0128-0

